# Impacts of the COVID-19 epidemic on the department of stomatology in a tertiary hospital: a case study in the General Hospital of the Central Theater Command, Wuhan, China

**DOI:** 10.1101/2020.09.11.20192450

**Authors:** Qingshan Dong, Angelica Kuria, Yanming Weng, Yu Liu, Yang Cao

## Abstract

**Objectives:** The aim of this study is to depict the impacts of COVID-19 pandemic on the clinical services and academic activities in the department of stomatology of a tertiary hospitals in Wuhan, China.

**Methods:** We obtained historical data of the Department of Stomatology from the Health Information System of the General Hospital of Central Theater Command, Wuhan, China between January 2018 and June 2020. Line plots were used to illustrate temporal trend of the variables. Mean ± standard deviation and median with interquartile range were used to summarize the variables. The Kruskal-Wallis equality-of-populations rank test was used to compare the difference between groups.

**Results:** A significant decrease was noted in the monthly average number of patients seeking the outpatient services for the year 2020. The monthly numbers of patients seeking outpatient services were decreased by two thirds from 2018 to 2020. The number of emergency cases also decreased significantly by 64% in 2020. The monthly number of teaching hours decreased from 3.8 ± 1.5 in 2018 and 4.7 ± 1.4 in 2019 to 1.7 ± 1.9 in 2020. The number of interns also decreased more than 70% in 2020.

**Conclusions:** The impacts of COVID 19 in the stomatology clinic were significant with notable decreases in clinical services and education offered to the stomatology students. We must find solutions to keep as many as needed dental profession stay on thriving and to remain on the frontline of healthcare.

## 1. Introduction

As a public health emergency of international concern, the severe acute respiratory syndrome coronavirus 2 (COVID-19) pandemic has resulted in over 23 million confirmed cases and 809 thousand deaths worldwide by August 23, 2020.[1, 2] According to the current evidence, COVID-19 virus is primarily transmitted between people through respiratory droplets and person-to-person contact routes.[3, 4] Dental patients who cough, sneeze, or receive dental treatment including the use of a high-speed handpiece or ultrasonic instruments make their secretions, saliva, or blood aerosolize to the surroundings.[5] Due to the characteristics of dental settings, the risk of cross infection may be high between dental practitioners and patients, and dental practitioners are at high risk of acquiring an infection while treating patients.[6] The impact of coronavirus on the dental community is eminent, the big challenge is how we can offer dental treatment despite the outbreak.[6, 7] Dental education programs and academic activities will also suffer from the ramifications of the pandemic.

The General Hospital of the Central Theater Command is a teaching hospital and one of the largest tertiary hospitals in Wuhan, where the COVID-19 epidemic broke in China. There are a total of 60 personnel in the Department of Stomatology of the hospital, including 34 stomatologists, 22 nurses, and 4 technicians. Fortunately, there were no personnel infected by COVID-19 during the epidemic in Wuhan because they strictly followed the Protocol for Prevention and Control of COVID-19 of the Chinese Center for Disease Control and Prevention throughout the epidemic.[8, 9] On 23 January 2020, the central government of China imposed a lockdown in Wuhan and other cities in Hubei province. Wuhan has been gradually reopened since April 1, 2020.[10]

The aim of this study was to depict the impacts of the COVID-19 epidemic on the clinical practices and academic activities of the department during January to June, 2020.

## 2. Materials and methods

We obtained historical data of the department from the Health Information System of the General Hospital of the Central Theater Command, Wuhan, China. The data were in monthly statistics and grouped into three broad categories (clinical services, education and academic activities/communication) between January 2018 and June 2020 to determine the impacts of COVID-19 on the department. Variables under clinical services included number of outpatient cases, emergency cases, operation cases, hospitalization and discharge of patients; numbers of interns and teaching hours were variables under education, while number of people attending scientific conference, lectures given by the personnel of the department, personnel study outside, and outside visitors to the department were variables for academic activities. Line plots were used to illustrate temporal trend of the variables during the study period. Mean ± standard deviation and median with interquartile range were used to summarize the variables. The Kruskal-Wallis equality-of-populations rank test was used to compare the difference between the medians of the variables during the same time period, i.e. from January to June, in 2018, 2019 and 2020. All the analyses were conducted in Stata 16.1 (StataCorp, College Station, Texas, USA).

There were no human subjects involved in the study, and only aggregated data were used in the analysis, and no private and confidential information could be disclosed, therefore the ethical approval is not applicable.

## 3. Results

### Clinical services

According to the results of this study, there was a decrease in the number of patients seeking clinical services in the stomatology clinic beginning January 2020. The month of February 2020 recorded almost zero patients in all variables for clinical services. Figure 1 shows a continuous trend of all the clinical services until the month of January 2020 when a decrease began. However, from the month of April 2020, an increase in the number of patients starts to be observed again. The monthly number of patients seeking the outpatient services during January and June was decreased by two thirds, which are 3561.3 ± 419.2 in 2018, 3996.2 ± 579.2 in 2019, and 1222.4 ± 1106.8 in 2020 (Table 1). The number of emergency cases attended to in this department also significantly decreased by 64%, from a monthly number of 125.2 ±17.8 in 2018 to 45.3 ± 18.6 in 2020. Table 1 compares the median numbers of all clinical services in the three years, and statistically significant differences are noted in all variables among the three years.

**Fig. 1.**
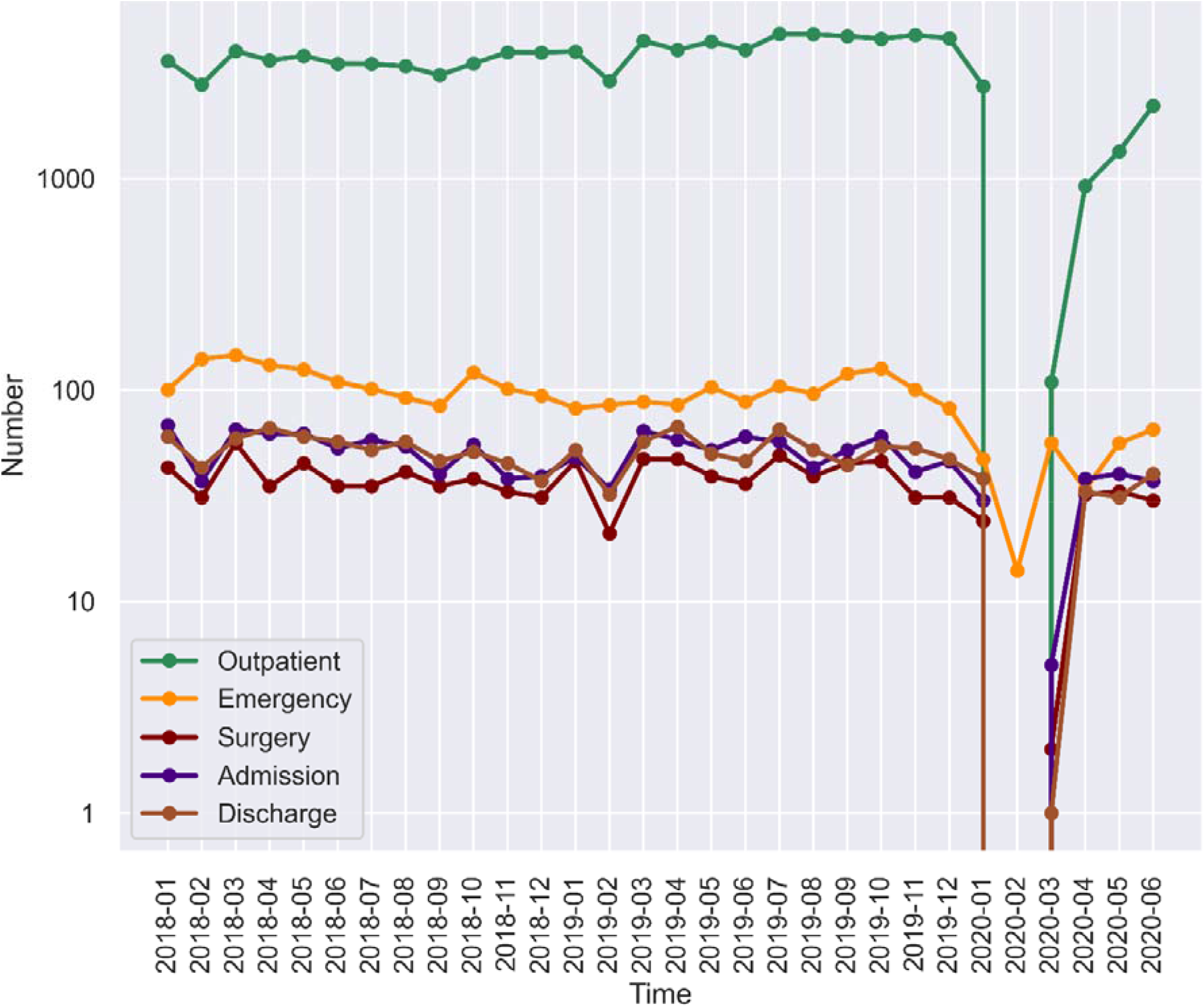
Time trend of the clinical services from January 2018 to June 2020

**Table 1.** Comparison of the clinical services, education, and academic activities (January - June)

| Variable | Mean (SD) |  |  | Median (Q1-Q3) |  |  | p-value* |
| --- | --- | --- | --- | --- | --- | --- | --- |
|  | 2018 | 2019 | 2020 | 2018 | 2019 | 2020 |  |
| Outpatient | 3561.3 (419.2) | 3996.2 (579.2) | 1222.5 (1106.8) | 3620.5 (3500.0-3812.0) | 4071.0 (3996.0-4450.0) | 1135.5 (109.0-2217.0) | 0.002 |
| Emergency | 125.2 (17.8) | 88.5 (7.4) | 45.3 (18.6) | 128.0 (109.0-140.0) | 86.5 (85.0-88.0) | 51.5 (34.0-56.0) | <0.001 |
| Operation | 40.8 (9.1) | 39.3 (10.1) | 20.2 (15.2) | 39.0 (35.0-45.0) | 42.5 (36.0-47.0) | 27.0 (2.0-32.0) | 0.019 |
| Admission | 57.8 (11.4) | 52.7 (10.8) | 25.0 (17.8) | 62.0 (53.0-65.0) | 55.0 (48.0-60.0) | 33.5 (5.0-38.0) | 0.011 |
| Dischagre | 57.5 (7.7) | 50.7 (11.7) | 23.8 (18.4) | 59.5 (57.0-60.0) | 51.0 (46.0-57.0) | 32.0 (1.0-38.0) | 0.005 |
| Interns | 20.5 (9.2) | 29.0 (10.1) | 5.7 (4.5) | 20.0 (18.0-30.0) | 31.0 (26.0-31.0) | 5.0 (3.0-8.0) | 0.005 |
| Teaching_hours | 3.8 (1.5) | 4.7 (1.4) | 1.7 (1.9) | 4.0 (4.0-5.0) | 5.0 (5.0-5.0) | 1.5 (0.0-2.0) | 0.029 |
| Conference_persons | 0.0 (0.0) | 0.7 (1.6) | 0.0 (0.0) | 0.0 (0.0-0.0) | 0.0 (0.0-0.0) | 0.0 (0.0-0.0) | 0.368 |
| Lectures | 0.0 (0.0) | 0.2 (0.4) | 0.0 (0.0) | 0.0 (0.0-0.0) | 0.0 (0.0-0.0) | 0.0 (0.0-0.0) | 0.368 |
| Study_outside | 1.0 (0.0) | 1.8 (0.4) | 1.3 (0.5) | 1.0 (1.0-1.0) | 2.0 (2.0-2.0) | 1.0 (1.0-2.0) | 0.015 |
| Visitors | 0.2 (0.4) | 0.2 (0.4) | 0.0 (0.0) | 0.0 (0.0-0.0) | 0.0 (0.0-0.0) | 0.0 (0.0-0.0) | 0.588 |

SD: standard deviation; Q1, the first quartile; Q3, the third Quartile
\* Kruskal-Wallis equality-of-populations rank test was used.

### Education

There is a decreasing trend for teaching hours and number of interns in the year 2020. This decrease was noted from the month of January 2020 (Figure 2). The monthly number of teaching hours decreased by > 50%, from 3.8 ± 1.5 in 2018 and 4.7 ± 1.4 in 2019 to 1.7 ± 1.9 in 2020. The number of interns also decreased by > 70%, from a monthly number of 20.5 ± 9.2 in 2018 and 29.0 ± 10.1 in 2019 to 5.7 ± 4.5 in 2020. Table 1 compares the medians of the education variables over the three years and shows statistically significant differences between the three years.

**Fig. 2.**
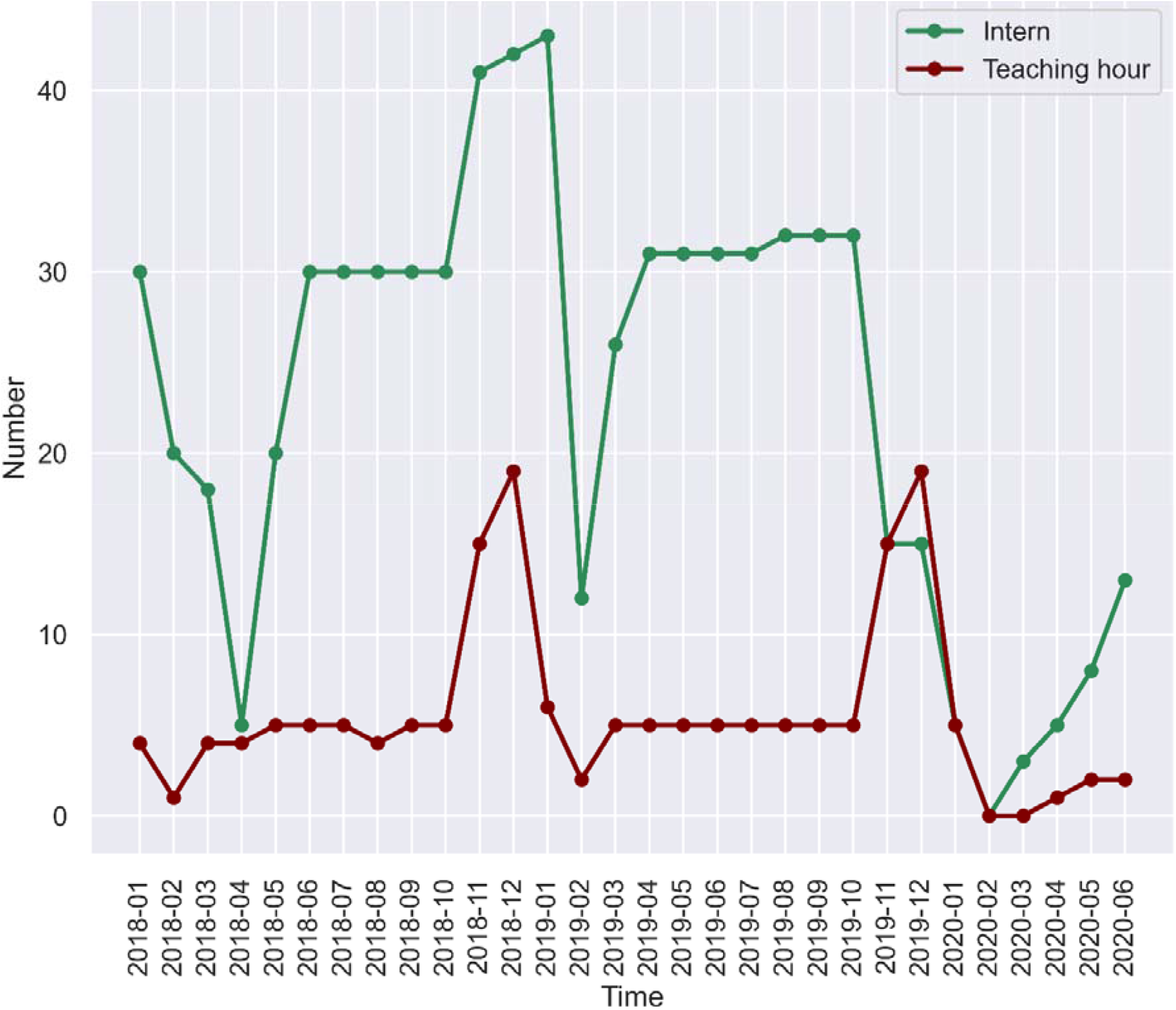
Time trend of the education from January 2018 to June 2020

### Academic Communication

Since academic activities/communication were usually taken in autumn and were much fewer in other seasons, there was no statistically difference noted in the time trend over the three years.

No statistically significant difference was found for academic activities/communication throughout the three years, except for the number of personnel studying outside (Table 1), however, the difference was no clinical significance.

## 4. Discussion

Stomatology clinics are among the departments in the health sector that are severely affected by infectious diseases mostly transmitted by respiratory droplets, and COVID-19 is no exception. This has been shown to be due to the nature of dental interventions and the proximity of the dental care provider to the oral region of the patient.[6, 11] Recent identification of COVID-19 in saliva [12] has also intensified the need for strict and effective infection control protocols for hospitals and dental practices in areas that are affected with COVID-19.

According to our study, all the clinical services and education in the stomatology clinic were significantly affected since the beginning of the epidemic in the city. The most significant impacts happened in the month of February, 2020 that recorded almost zero patients in the number of outpatients, emergency services, operations and hospitalization. Although the decline in the number of patients seeking services was expected after the emergence of the epidemic due to fear and recommendations from the government of China to stay indoors [6, 10], reports of the first human to human transmission seems to have also severely affected these services as this was first reported in late January 2020.[13] Our study collaborating with other studies show that in times of the similar epidemics happening the clinical services in stomatology clinics having been severely affected.[6, 11, 14]

The education in this department was affected with a decline in the number of teaching hours and the number of interns compared to other years. Although practical sessions are the preferred format for the department, online sessions have also shown promising results as reported by Liu et al.[15]

There was no impact observed on academic activities so far due to their inherent seasonal pattern. However, the long-term effects still need to be investigated in the future.

## 5. Conclusion

The impacts of COVID 19 on the stomatology clinic are significant with a decrease in clinical services and education offered to the stomatology students. To mitigate the negative impacts of the epidemic on the stomatology clinics, it is necessary to review the guidelines and protocols of the clinics, and encourage people seek for dental services with enough protection against COVID-19, therefore to keep as many as needed dental profession stay on thriving and remain on the frontline of healthcare.

## Data Availability

Data available on request from the first author.

## Data Availability

Data available on request from the first author.

